# Genetic architecture of immune-mediated thrombotic thrombocytopenic purpura in Han Chinese individuals

**DOI:** 10.64898/2026.09.21.26363596

**Authors:** Zeyu Gan, Jingjing Jiang, Zilin Lai, Lv Xiong, Yajie Ding, Shanshan Luo, Yu Hu, Xingjie Hao, Jun Deng

**Author notes:** Corresponding author: Jun Deng,; Xingjie Hao,; Yu Hu, and Shanshan Luo,. These authors contributed equally to this work (Zeyu Gan, Jingjing Jiang).

## Abstract

Thrombotic thrombocytopenic purpura (TTP) is a rare, life-threatening thrombotic microangiopathy caused by a severe deficiency of the ADAMTS13 protease. We performed the first large-scale genome-wide association study (GWAS) of TTP in 1,344 individuals of Han Chinese ancestry, comprising a discovery cohort (n = 913) and a validation cohort (n = 431). We identified two independent susceptibility loci, including the known locus 6p21 and the novel locus 14q32. At the known locus 6p21, the lead signal rs2187668 is near HLA-DQA1 and strongly associated with TTP risk (*P* = 5.35 × 10^-10^; Odds Ratio [OR] = 3.94). At the novel locus 14q32, the lead SNP rs1024350 (*P* = 1.01 × 10^-10^; OR = 1.90) is located between LINC00221 and MIR5195. Fine-mapping prioritized rs2187668 as the leading signal at 6p21, whereas HLA imputation and stepwise conditional analysis further pinpointed amino acid position 74 of HLA-DRB1 as the primary driver of the broader MHC association (*P* = 2.28 × 10⁻¹⁰), superseding classical allele-based models. Structural analysis revealed that position 74 lines pocket 4 of the HLA-DRB1 peptide-binding groove; the risk-conferring Leu/Arg residues at this position carried by HLA-DRB1*08:03 and DRB1*03 may favour presentation of ADAMTS13-derived autoantigenic peptides, whereas the common Ala residue is protective. In summary, we reconstructed the genetic architecture of TTP in East Asians and identified structural gatekeepers of autoimmunity, providing novel targets for precision medicine and ethnic-specific risk stratification.

## Introduction

Thrombotic thrombocytopenic purpura (TTP) is a rare, life-threatening thrombotic microangiopathy. It is clinically characterized by a pentad of symptoms, including acute thrombocytopenia, microangiopathic hemolytic anemia, and multiorgan ischemia— most notably affecting the neurological and renal systems^1^. The global annual incidence is estimated at 2 to 6 cases per million, with a predilection for women aged 40–50 years^2^. The pathogenesis of TTP centers on a severe deficiency of the von Willebrand factor (VWF)-cleaving protease, ADAMTS13 (activity <10%). While congenital TTP results from rare biallelic mutations in the ADAMTS13 gene on chromosome 9^3^, approximately 95% of cases are immune-mediated TTP, triggered by inhibitory autoantibodies that suppress enzyme activity or accelerate its clearance^4^. This defect impairs the proteolytic cleavage of ultra-large VWF multimers, leading to the formation of platelet-rich microthrombi and a high risk of recurrence^5,6^.

Notably, previous genetic studies have identified key susceptibility loci within the major histocompatibility complex (MHC). In European populations, targeted immunochip analyses identified significant associations centered on HLA-DRB1 alleles^7^. Specifically, the HLA variant rs6903608 has been robustly linked to disease onset and relapse in Caucasian cohorts^2^. Furthermore, the HLA class II locus, particularly the DRB1*11 and DQB1*03 alleles, has been consistently associated with a loss of immune tolerance to ADAMTS13, contributing to the autoimmune etiology of TTP^8^. Complementing these findings, research in Japanese populations has implicated distinct HLA susceptibility loci, suggesting a mechanistic role for shared ADAMTS13-derived peptides that bind to specific HLA-DR alleles to trigger autoantibody production^9^. Beyond the MHC region, a European GWAS recently identified a novel genetic locus on chromosome 3 associated with TTP^10^. However, current genetic research remains largely confined to Caucasian populations, with extremely limited data available for East Asians. To date, no large-scale GWAS has been conducted in non-Caucasian groups, leaving a critical gap in our understanding of TTP etiology in populations where HLA allele frequencies and linkage disequilibrium (LD) structures differ significantly. Elucidating genome-wide genetic features is therefore crucial for identifying populations at higher risk for disease onset and recurrence.

In this study, we present the first GWAS of TTP in Han Chinese population, featuring the largest Asian cohort reported to date. By integrating two-stage association testing, Sum of Single Effects (SuSiE)-based fine-mapping, and high-resolution HLA imputation, we further elucidate the genetic architecture within the HLA region, providing new insights into the molecular basis of TTP.

## Method

### Study population and sample collection

The study was conducted in two stages: a discovery stage and a validation stage. The discovery cohort consisted of 913 Han Chinese individuals (172 cases and 741 controls), while the validation cohort included 431 individuals (129 cases and 302 controls). All thrombotic thrombocytopenic purpura (TTP) cases were diagnosed based on clinical presentation of thrombosis and hemostasis group, Chinese society of hematology and Chinese medical association^11,12^. And a documented ADAMTS13 activity levels below 10% concomitant with positive ADAMTS13 inhibitors. Controls were healthy individuals with no history of TTP.

The study design is illustrated in **Figure 1**. All the patients with TTP in this study were recruited from China mainland, and all patients were Han Chinese. The study was approved by the Ethical Board of Tongji Medical College, Huazhong University of Science and Technology, China (Ethics code: UHCT-IEC-SOP-016-03-01), and written informed consent was obtained from all participants.

**Figure 1.**
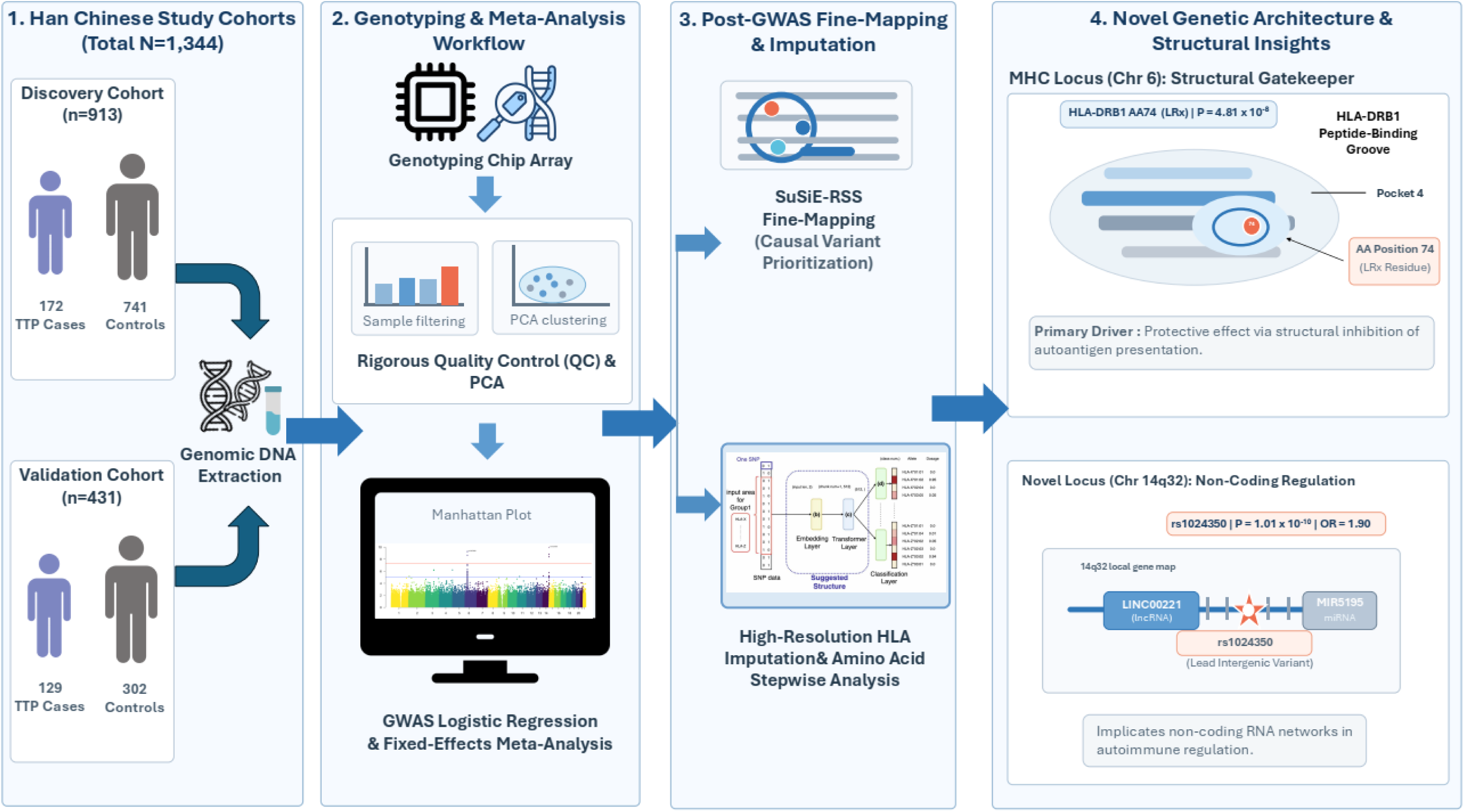
Overview of the study design and analytical workflow. The study included discovery and validation cohorts of Han Chinese individuals, followed by genotyping, sample and variant quality control, population-structure assessment, genome-wide association testing, fixed-effects meta-analysis, fine-mapping, HLA imputation, amino-acid association analysis, and structural interpretation of the identified susceptibility signals.

### Genotyping and quality control

Genotyping was performed using the Illumina Infinium Asian Screening Array. Stringent quality control (QC) was applied to both samples and single nucleotide polymorphisms (SNPs) using PLINK v2.0^13^ and R-4.5.0. For SNPs, we removed duplicated variants sharing identical chromosomal positions and variants with missing alleles. Autosomal variants were then filtered by minor allele frequency (MAF < 0.01), Hardy–Weinberg equilibrium test (*P* < 1 × 10^-6^ using mid-*P* adjustment), and SNP missingness (GENO > 0.05). Samples were excluded if they had: (i) a call rate < 95%; (ii) high missingness (MIND > 0.10); (iii) extreme heterozygosity and inbreeding coefficients (F Coefficient outside −0.1 to 0.3); or (iv) cryptic relatedness (kinship coefficient > 0.0884). Following QC, principal component analysis was performed to capture population structure.

### Genome-wide association analysis

All genome-wide association analyses were conducted using logistic regression implemented in PLINK v2.0. To account for potential population stratification, the top five principal components (PCs), along with age (only in validation cohort) and sex, were included as covariates. Manhattan plots and quantile–quantile (Q–Q) plots were generated using the qqman (v0.1.9) package^14^. The threshold for genome-wide significance was set at *P* < 5 × 10^−8^.

### Meta-analysis

A fixed-effects meta-analysis was performed using METAL (v2020-05-05)^15^ to combine results from the discovery and validation stages, totaling 1,344 individuals. Heterogeneity between cohorts was assessed using Cochran’s Q test and the *I*^2^ statistic. To identify independent signals, we performed LD-based clumping (*r*^2^ < 0.1 within a 1000 kb window) based on the study genotype data, using UCSC reference sequence hg19 gene range lists for annotation. The primary significance threshold for clumping was set at *P* < 1 × 10^-5^.

### Fine-mapping of susceptibility loci

To meticulously dissect the genetic architecture and pinpoint the potential causal variants of TTP within complex regions, we performed a targeted fine-mapping strategy using the Sum of Single Effects (SuSiE-RSS) model^16,17^ via its R package ’susieR’ (v0.14.2)^17,18^. Meta-analysis summary statistics were integrated with a reference LD matrix, following rigorous allele alignment and *β* flipping. The model was executed with a maximum of 5 causal effects (L=5). Potential causal variants were prioritized by calculating posterior inclusion probabilities (PIP) and 95% credible sets (CS). To ensure signal reliability, a purity filter was applied, retaining only CS with a minimum absolute correlation (|r|) of 0.5 among member variants. For the MHC region on chromosome 6, where LD is extended and differs between cohorts, fine-mapping the meta-analysis against a single pooled reference panel was unreliable; we therefore performed cross-cohort fine-mapping with SuSiEx^19^, supplying each cohort’s GWAS summary statistics together with its own in-sample LD reference.

### HLA imputation and association analysis

To further elucidate the role of HLA alleles in TTP susceptibility, MHC region variants (chr6: 25–34 Mb) and classical HLA alleles were imputed using the Michigan Imputation Server 2 with the four-digit multi-ethnic HLA reference panel v2^20^, and we filtered those low quality variants (*R*^2^ < 0.7) after imputation.

Association analysis for HLA features and amino acid residues was performed using logistic regression, adjusted for age (only in validation cohort), sex, and the top five PCs. To identify the primary driver of the MHC signal, we conducted stepwise conditional analysis by adjusting for the lead amino acid residue (HLA-DRB1 position 74). The LD structure between the lead amino acid residues and classical HLA alleles was calculated using *r*^2^. To decouple the effects of the amino acid residue from its host allele, we performed haplotype-based risk stratification and interaction analyses. Odds ratio (OR) and 95% confidence interval (CI) were calculated for individuals categorized by their carrier status of the risk amino acid and the associated classical allele. Genomic control was not applied to the MHC meta-analysis, because within this densely associated region the inflation statistic (λ ≈ 1.5) reflects genuine HLA signal in LD rather than stratification, which was already controlled by principal components. Amino-acid residues at position 74 (Ala/Glu/Leu/Gln/Arg) were taken from the imputed amino-acid markers.

HLA-DRB1∼DQB1 haplotypes were reconstructed from the imputed two-field classical alleles using an expectation–maximization algorithm, and haplotype associations were tested by covariate-adjusted logistic regression^21^. The relative contributions of HLA-DRB1 and HLA-DQB1 were assessed by reciprocal conditional omnibus likelihood-ratio tests.

## Results

### Study overview and quality control

We conducted a two-stage genome-wide association study (GWAS) to investigate the genetic basis of TTP in the Han Chinese population. The overall study design and analytical workflow are summarized in **Figure 1**. The discovery stage comprised a cohort of 913 individuals, including 172 TTP cases and 741 healthy controls. To confirm the findings from the discovery stage, an independent validation cohort of 431 individuals (129 cases and 302 controls) was analyzed. Detailed demographic and clinical characteristics for both cohorts are presented in **Table S1**.

Prior to the formal quality control process, we conducted a comprehensive descriptive analysis of the raw genotype data for both the discovery and validation cohort (**Supplementary Figures S1** and **Supplementary Figures S2**). After stringent QC, we retained 524,866 and 510,182 high-quality autosomal SNPs from the discovery and validation stages, respectively.

### Genome-wide association identifies loci associated with TTP

We performed a genome-wide meta-analysis combining the discovery (n=913) and validation (n=431) cohorts. The genomic inflation factors ( *λ* ) were 1.041 for the discovery stage, 1.029 for the validation stage (**Supplementary Figures S3-S4**), and 1.024 for the meta-analysis (**Figure 2**), indicating that population stratification was well-controlled and had a minimal impact on the results. The meta-analysis identified two independent susceptibility loci reaching genome-wide significance (*P* < 5 × 10^-8^) or suggestive significance with robust evidence across both stages (**Figure 2**), including two signals at 6p21 and one signal at 14q32 (**Table 1**).

**Figure 2.**
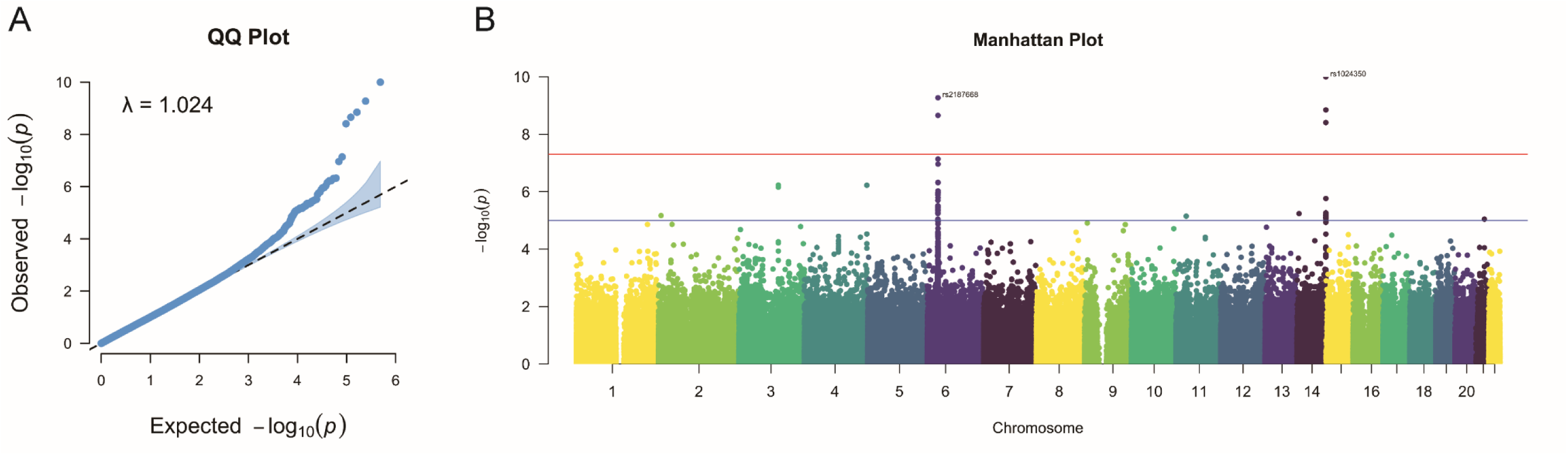
GWAS meta-analysis of thrombotic thrombocytopenic purpura. (A) Quantile-quantile plot of observed versus expected association *P* values in the meta-analysis. (B) Manhattan plot of genome-wide association results across autosomes. The red line indicates the genome-wide significance threshold of *P* = 5 × 10^−8^, and the blue line indicates the suggestive significance threshold of *P* = 1 × 10^−5.^

**Table 1.** Genome-wide significant variant associated with thrombotic thrombocytopenic purpura.

| rsID | Chr | Position<br>(Hg19) | Risk Allele | Non-risk Allele | Stage | Risk allele frequency |  | OR (95% CI) | P-value |
| --- | --- | --- | --- | --- | --- | --- | --- | --- | --- |
|  |  |  |  |  |  | TTP | Control |  |  |
| rs2187668 | 6 | 32605884 | A | G | Discovery | 0.124 | 0.061 | 7.13 (3.43-14.81) | 1.40×10<sup>-7</sup> |
|  |  |  |  |  | Validation | 0.105 | 0.064 | 2.86 (1.67-4.90) | 0.0001 |
|  |  |  |  |  | Meta-Analysis | 0.113 | 0.063 | 3.94 (2.56-6.08) | 5.35×10<sup>-10</sup> |
| rs6906021 | 6 | 32626311 | T | C | Discovery | 0.344 | 0.452 | 0.48 (0.33-0.69) | 6.52×10<sup>-5</sup> |
|  |  |  |  |  | Validation | 0.369 | 0.519 | 0.53 (0.40-0.70) | 7.51×10<sup>-6</sup> |
|  |  |  |  |  | Meta-Analysis | 0.358 | 0.5 | 0.51 (0.41-0.64) | 2.21×10<sup>-9</sup> |
| rs1024350 | 14 | 107141122 | G | A | Discovery | 0.581 | 0.457 | 1.89 (1.35-2.63) | 0.0002 |
|  |  |  |  |  | Validation | 0.581 | 0.414 | 1.90 (1.50-2.40) | 1.32×10<sup>-7</sup> |
|  |  |  |  |  | Meta-Analysis | 0.581 | 0.427 | 1.90 (1.56-2.30) | 1.01×10<sup>-10</sup> |

Positions are based on GRCh37/hg19. The tested (risk) allele is the effect allele in the association model; odds ratios and 95% confidence intervals are reported for this allele (OR > 1 indicates increased susceptibility). Risk-allele frequencies are shown separately for TTP cases and controls. Chr, chromosome; CI, confidence interval; OR, odds ratio; TTP, thrombotic thrombocytopenic purpura.

The most prominent association signal rs2187668 at 6p21(**Figure 3A**) is located in the intronic region of HLA-DQA1, this SNP emerged as a leading signal. In the meta-analysis, the risk allele A was strongly associated with TTP (*P* = 5.35 × 10^-10^), exhibiting a substantial effect size with an odds ratio (OR) of 3.94 (95% CI: 2.56 - 6.08). The other signal rs6906021 at 6p21 (**Figure 3B**) is located at the downstream of HLA-DQB1, this variant also showed significant association in the meta-analysis (*P* = 2.21 × 10^-9^), with the T allele conferring a protective effect (OR = 0.51, 95% CI: 0.41 - 0.64). The consistency of effect directions and significance levels across both the discovery and validation stages underscores the robust role of the HLA-DQ subregion in TTP susceptibility among Han Chinese.

**Figure 3.**
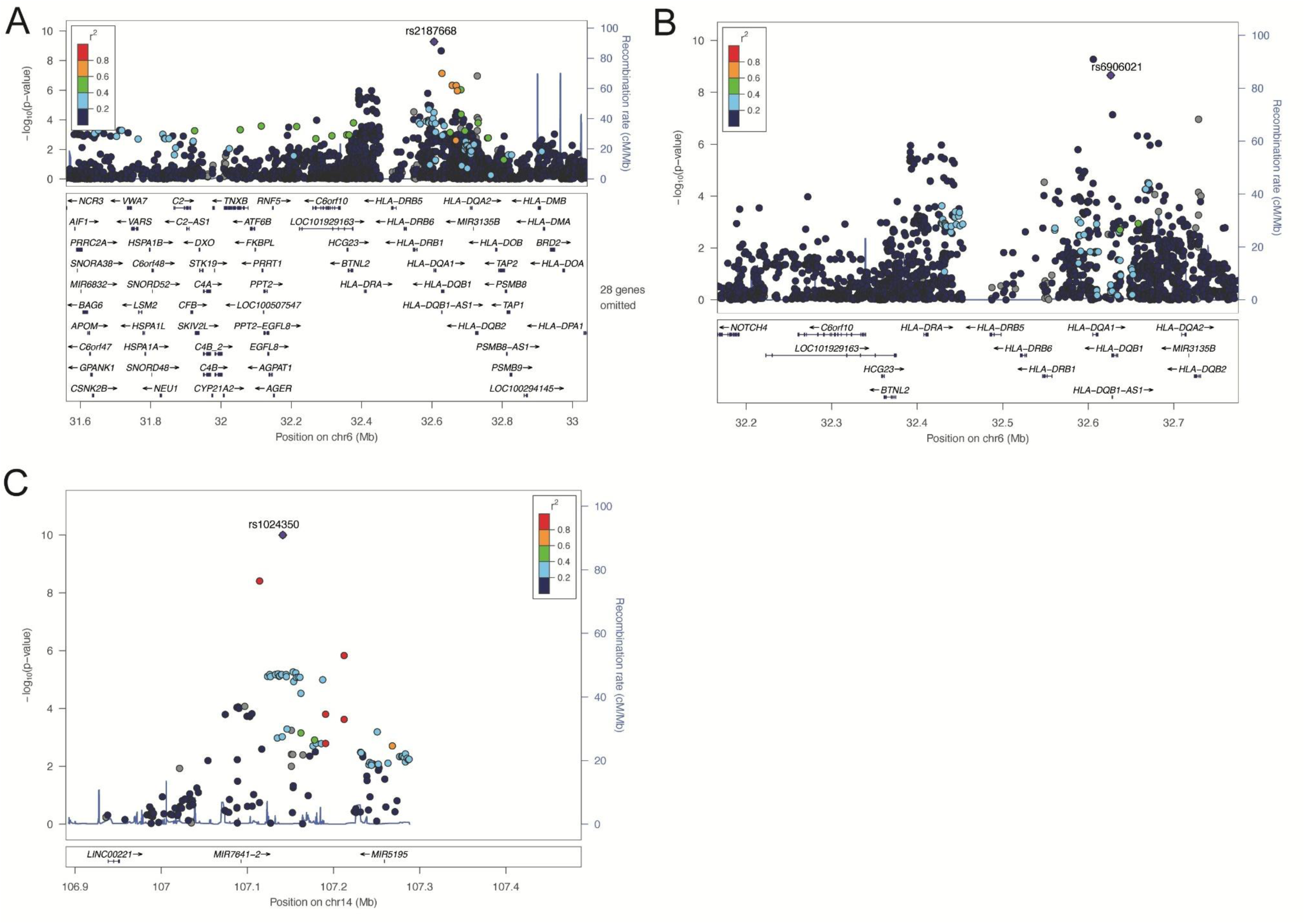
Regional association plots for the three lead signals identified in the GWAS meta-analysis. (A) Regional association plot for rs6906021 at 6p21. (B) Regional association plot for rs2187668 at 6p21. (C) Regional association plot for rs1024350 at 14q32. Genomic positions are based on GRCh37/hg19. Points are colored according to linkage disequilibrium with the lead variant, and recombination rates are shown in blue.

Beyond the 6p21 region, we identified a novel susceptibility locus on chromosome 14 (**Figure 3**). The lead SNP rs1024350 is located between LINC00221 and MIR5195. It reached genome-wide significance in the meta-analysis (*P* = 1.01 × 10^-10^), with the ’G’ allele significantly increasing TTP risk (OR = 1.90, 95% CI: 1.56 - 2.30). Notably, this locus showed highly consistent effect sizes in both the discovery (OR = 1.89) and validation (OR = 1.90) stages, highlighting it as a reliable novel genetic marker for TTP outside the classical HLA region.

Other suggestive signals, including an intronic variant in TIMMDC1 on chromosome 3 and an intergenic variant near STXBP6/NOVA1 on chromosome 14 were also observed (**Table S2, Figure S7**), providing further candidates for future functional validation.

### Fine-mapping pinpoints potential causal variants at associated loci

To resolve the causal architecture underlying the observed GWAS signals, we performed fine-mapping using the SuSiE-RSS model. Because the MHC harbours extended, cohort-specific LD that a pooled reference panel does not capture, we fine-mapped 6p21 with SuSiEx using each cohort’s in-sample LD. This resolved two independent credible sets: one led by rs2187668 (chr6:32.61 Mb; PIP = 0.999, supported in both cohorts) and one at chr6:32.39 Mb (lead rs114188689 near HLA-DRA, driven by the discovery cohort). Although LD clumping had flagged rs2187668 and rs6906021 as the two lead variants at 6p21, cross-cohort fine-mapping showed that rs6906021, despite reaching genome-wide significance in the meta-analysis, did not enter any credible set (PIP ≈ 0), indicating that its marginal association tags the underlying HLA signal rather than an independent causal SNP. The two credible sets localized to different variants in the two cohorts, reflecting the complex regional structure of the MHC (**Table S6, Figure S8**).

For 14q32 region, fine-mapping narrowed the association signal to a highly restricted genomic interval. The GWAS lead SNP rs1024350 was confirmed as the most likely causal variant with a PIP of 0.805, while a second variant, GSA-rs7157975, was included in the same credible set with of PIP = 0.155. These two variants were in strong LD (*r*^2^ = 0.913) and demonstrated high purity, indicating that these two variants are in strong LD and together account for over 95% of the posterior probability at this locus (**Table S3, Figure S9**). Given its superior PIP and status as the lead meta-analysis SNP, rs1024350 remains the top functional candidate for further investigation at 14q32.

### Fine-mapping of the MHC locus by genotype imputation

To pinpoint the causal variants driving the genetic association of TTP within the MHC, we performed HLA imputation followed by a comprehensive meta-analysis of two cohorts. The strongest association signal was localized to the HLA-DRB1 gene, with the most significant variant identified as the amino acid residue at HLA-DRB1 position 74 (*P* = 2.28 × 10^-10^, **Figure 4**). To determine whether HLA-DRB1 position 74 fully accounts for the observed MHC association, we conducted a stepwise conditional analysis. Conditioning on the dosage of the position-74 signal resulted in a near-complete abolition of the association signals across the entire MHC region. For instance, the significance of the lead GWAS SNP rs6906021 dropped to *P* = 4.18 × 10⁻⁵ and no marker in the MHC reached genome-wide significance (*P* < 5 × 10⁻⁸), indicating a single independent signal (**Figure S5** and **Table S4**).

**Figure 4.**
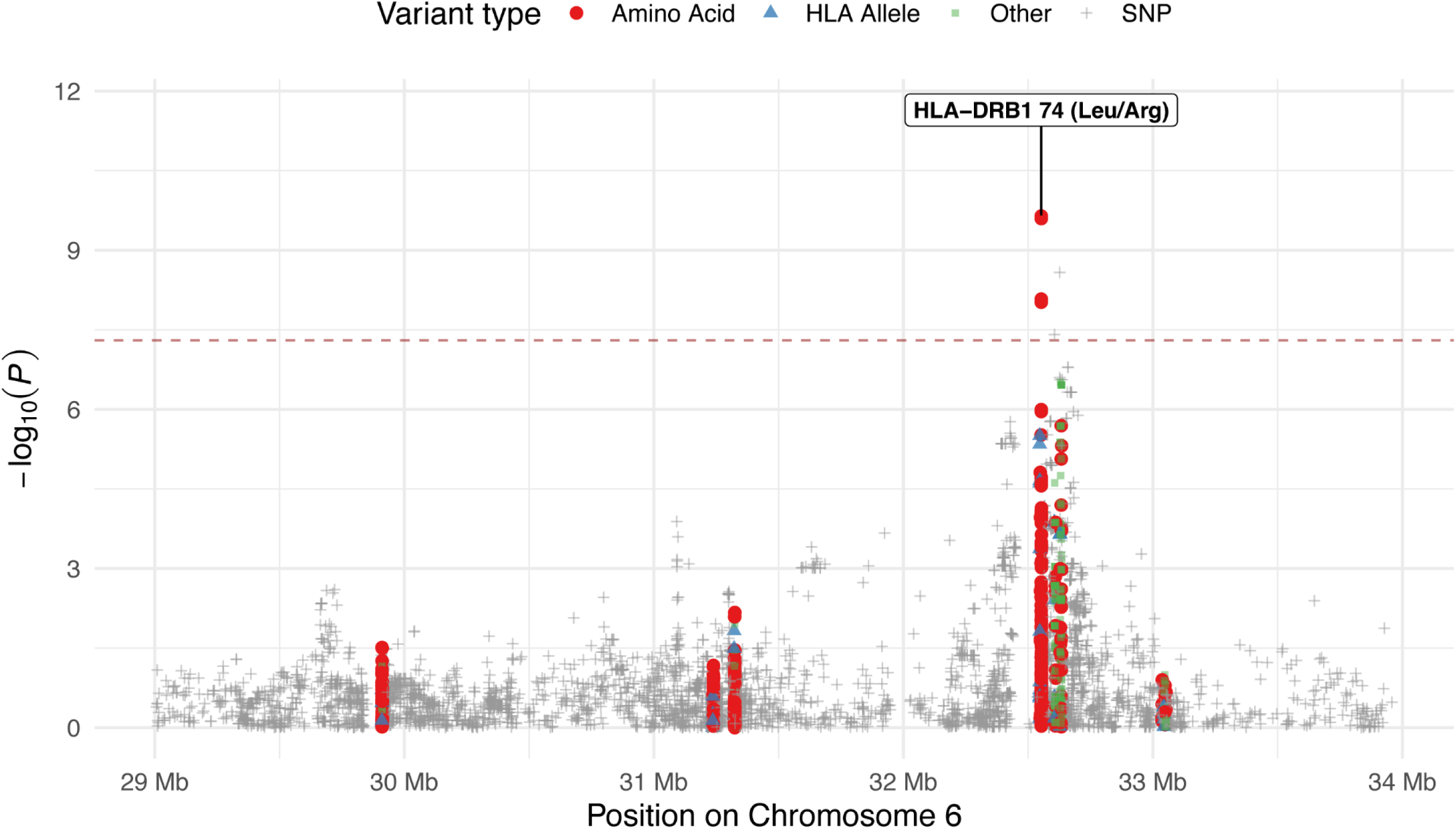
Regional association analysis of imputed variants at the MHC region. Regional association results for imputed SNPs, classical HLA alleles, and HLA amino-acid variants across the MHC region are shown. Variant classes are indicated by color and shape. The lead signal, HLA-DRB1 amino-acid position 74 (Leu/Arg), is labeled. The horizontal dashed line indicates the genome-wide significance threshold of *P* = 5 × 10^−8^.

Furthermore, we explored the LD structure between HLA-DRB1 position 74 and classical HLA alleles. The risk residue 74-Leu was strongly tagged by HLA-DRB1*08:03 (*r*² = 0.88) and 74-Arg by HLA-DRB1*03 (*r*² ≈ 1.0), while both risk residues also appeared on additional HLA backgrounds, indicating a cross-haplotype, residue-level effect (**Figure S6**). Notably, no single classical DRB1 allele reached genome-wide significance, whereas the position-74 residue did, confirming that the amino-acid classification supersedes allele-based models. Carriage of 74-Leu or 74-Arg conferred an increased risk of TTP that was consistent across both cohorts (OR = 2.94 in the discovery; OR = 2.02 in the validation; OR_meta_ = 2.55, 95% CI: 1.91 - 3.41, *P* = 2.28 × 10⁻¹⁰; **Table 2**); equivalently, the common non-Leu/Arg configuration was protective (OR_meta_ = 0.39, 95% CI: 0.29 - 0.52). The effect was independent of HLA-DRB1*08:03 carrier status, and the amino acid × allele interaction was non-significant (*P*_interaction_ > 0.05; **Table S5**). Structurally, position 74 lines pocket 4 of the HLA-DRB1 peptide-binding groove; the risk-associated Leu/Arg residues may alter the pocket’s physicochemical properties to favour presentation of ADAMTS13-derived autoantigenic peptides, whereas the common Ala residue is protective.

**Table 2.** Association of HLA-DRB1 amino-acid position 74 with thrombotic thrombocytopenic purpura.

| Marker (AA state tested) | Chr | Position<br>(Hg19) | Stage | Risk allele frequency |  | OR (95% CI) | P-value |
| --- | --- | --- | --- | --- | --- | --- | --- |
|  |  |  |  | TTP | Control |  |  |
| HLA-DRB1 position 74 (exon2): Leu/Arg | 6 | 32552669 | Discovery | 0.211 | 0.108 | 2.94 (2.04-4.17) | 7.93×10<sup>-9</sup> |
|  |  |  | Validation | 0.191 | 0.115 | 2.02 (1.25-3.23) | 3.64×10<sup>-3</sup> |
|  |  |  | Meta-Analysis | 0.202 | 0.110 | 2.55 (1.91-3.41) | 2.28×10<sup>-10</sup> |

Positions are based on GRCh37/hg19. The risk allele is the allele tested in the association model; odds ratios and 95% confidence intervals are reported for the risk allele. Risk allele frequency are shown separately for TTP cases and controls. Chr, chromosome; CI, confidence interval; OR, odds ratio; TTP, thrombotic thrombocytopenic purpura.

To place the MHC association in the context of HLA class II haplotypes, we reconstructed HLA-DRB1∼DQB1 haplotypes and tested their association with TTP. Two haplotypes conferred increased risk: DRB1*03:01–DQB1*02:01 (DR3/DQ2; OR = 4.16, 95% CI: 2.18–7.97, *P* = 1.7 × 10⁻⁵) and DRB1*08:03–DQB1*06:01 (OR = 2.60, 95% CI: 1.56–4.32, *P* = 2.4 × 10⁻⁴; **Table S7**, **Figure S10**). The latter is the risk haplotype reported in Japanese immune-mediated TTP^9^, indicating a shared HLA class II susceptibility haplotype across East Asian populations. The two independent SNP credible sets mapped to distinct DR–DQ haplotypes: rs2187668 tagged DR3/DQ2 (*r*² = 0.86 with DRB1*03:01), whereas rs6906021 tagged the HLA-DQB1*03 region (*r*² = 0.22 with DQB1*03:03). Consistently, the rs6906021 association remained after conditioning on HLA-DRB1 position 74 (*P* = 4.2 × 10⁻⁵), was abolished by additionally conditioning on HLA-DQB1 (*P* = 0.65), and persisted after conditioning on the full HLA-DRB1 allele (*P* = 1.1 × 10⁻⁴), suggesting an HLA-DQ contribution beyond DRB1 position 74. However, because HLA-DRB1 and HLA-DQB1 are in very strong LD in this region, neither gene reached significance conditional on the other (*P* = 0.065– 0.067), and the DR and DQ contributions could not be cleanly separated in the present sample (**Table S8**).

## Discussion

Our study addresses a longstanding disparity in the genomic understanding of TTP, which has historically been derived almost exclusively from Caucasian cohorts. By conducting the largest GWAS of TTP in the Han Chinese population to date. we elucidate that TTP susceptibility is not merely driven by distinct HLA alleles, but is fundamentally orchestrated by specific structural properties of the HLA-DRB1 peptide-binding groove, specifically the amino acid residue at position 74. Furthermore, we identify a novel locus at 14q32 involving the long non-coding RNA LINC00221, expanding the pathological framework of TTP from protein-coding variants to the complex landscape of non-coding RNA-mediated immunoregulation.

The most defining feature of our data is the pinpointing of HLA-DRB1 amino acid position 74 as the primary independent driver of major histocompatiblity complex (MHC)-associated risk, a finding that refines the classical allele-based risk models. While previous studies in European populations have robustly linked risk to the HLA-DRB1*11 and HLA-DQB1*03 haplotypes^22–25^, our HLA imputation and conditional analyses indicate that, in the Han Chinese population, the broader MHC association resolves to a specific residue variation at position 74. This position is critical because it lines pocket 4 of the HLA-DRB1 antigen-binding groove, a site that governs the anchoring of peptide side chains^26^. We postulate that the risk-associated Leu/Arg residues at position 74 carried by HLA-DRB1*08:03 and DRB1*03:01, confer a pocket 4 configuration that stabilizes binding of the immunodominant ADAMTS13 C-CUB peptide^27,28^ , promoting formation of the MHC–peptide–TCR complex required to break T-cell tolerance^29^. Conversely, the common Ala residue destabilizes this interaction and is protective, providing a mechanistic explanation for the observed effect. This finding underscores a phenomenon of structural convergence in TTP pathogenesis: while European cohorts implicate DRB1*11, our data identify DRB1*08:03 (encoding Leu at position 74) as a risk allele in Han Chinese, consistent with HLA-DRB1*08:03 as a susceptibility allele in Japanese immune-mediated TTP^9^. At the haplotype level, DRB1*08:03–DQB1*06:01 itself was a risk haplotype in our cohorts, reinforcing a shared HLA class II susceptibility across East Asians and echoing the amino-acid-level convergence. This DR–DQ haplotype is implicated in Japanese TTP. This cross-ancestry pattern, in which distinct risk alleles converge on the physicochemical properties of the peptide-binding pocket^30^, points to a universal mechanism of autoimmunity that could be targeted therapeutically.

Beyond the MHC, the discovery of the 14q32 locus (rs1024350) represents a paradigm shift, implicating the non-coding genome in TTP etiology. This variant resides in the intergenic region between LINC00221 and MIR5195. Although LINC00221 has largely been characterized in the context of neoplastic progression^31,32^, emerging transcriptomic evidence suggests it functions as a competing endogenous RNA (ceRNA) that modulates immune cell infiltration and cytokine signaling^33,34^. It is biologically plausible that the risk allele at rs1024350 alters the expression or secondary structure of LINC00221, disrupting a local ceRNA network that normally restrains B-cell hyperactivation^32,34^. Intriguingly, previous phenome-wide association studies have linked this specific locus to variations in antibody responses against viral antigens and the usage of the IGHV1-69 gene segment^35,36^. This connection is particularly provocative given the two-hit hypothesis of TTP, where infection often precipitates acute episodes^37^. We speculate that rs1024350 may bias the naive B-cell repertoire toward IGHV1-69^38,39^, facilitating the generation of polyreactive antibodies that cross-react with ADAMTS13 upon viral challenge^40,41^. Thus, this locus may represent the genetic bridge between environmental triggers and autoimmune onset.

Our analysis of chromosome 3 provides a compelling example of how trans-ethnic studies can resolve complex LD blocks. In European cohorts, susceptibility signals in this region were attributed to POGLUT1^10^, a glycosyltransferase modifying ADAMTS13^42^. While our Chinese cohort replicated the signal in this region, the peak association mapped more closely to TIMMDC1, a gene encoding a mitochondrial complex assembly factor^43^. This discrepancy likely reflects differences in LD structure between Han Chinese and European populations rather than distinct biological mechanisms. However, the involvement of TIMMDC1 cannot be discounted since mitochondrial dysfunction in endothelial cells is a known driver of pro-thrombotic phenotypes^44,45^. It is possible that in the Han Chinese genetic background, variants in TIMMDC1 compromise endothelial resilience to oxidative stress^46,47^, thereby lowering the threshold for microvascular thrombosis in the presence of anti-ADAMTS13 antibodies^48–50^. This suggests that the chromosome 3 locus may exert a dual-hit effect impairing both enzymatic glycosylation (POGLUT1) and endothelial metabolic fitness (TIMMDC1).

These findings must be interpreted within the context of certain limitations. Although this represents the largest TTP GWAS in an Asian population, the sample size remains modest compared to studies of common diseases, potentially limiting our power to detect rare variants with smaller effect sizes. And our study cohort was restricted to patients with ADAMTS13 activity levels below 10% concomitant with positive ADAMTS13 inhibitors. Furthermore, while our finding of 14q32 loci is very significant and robust fine-mapping methods is applied, the loci also need to be evaluated in other ancestries to evaluate the generalizability, and the precise molecular mechanism by which LINC00221 influences TTP susceptibility requires experimental validation in relevant cell models. In addition, the strong LD between HLA-DRB1 and HLA-DQB1, together with the modest number of cases, precluded a definitive separation of DR- and DQ-specific effects; the suggestive HLA-DQ contribution will require larger cohorts or informative recombinant haplotypes to confirm. However, we conducted a multi-stage study design to minimize false-positive findings while maximizing power and efficiency. And the high replication consistency across the two stages and the biological coherence of the identified loci provide strong confidence for these results.

In summary, this study reconstructs the genetic architecture of TTP in the Han Chinese population, moving beyond simple allele associations to implicate specific amino acid residues and non-coding regulatory networks. The identification of HLA-DRB1 position 74 as a structural gatekeeper of autoimmunity and the discovery of the LINC00221 locus offer novel targets for precision medicine. Clinically, these markers could be integrated into polygenic risk scores to identify patients at high risk of relapse or to stratify carriers who may require closer monitoring during infectious episodes. Ultimately, our work highlights the necessity of non-European genomic data in completing the global puzzle of complex autoimmune diseases.

## Data Availability

Supplementary information about this paper is available online. Further inquiries can be directed to the corresponding author X.H.

## Acknowledgments

The computation is completed in the HPC Platform of Huazhong University of Science and Technology.

## Contributions

YH, XJH, and SSL conceived the idea for the study, LX, YJD performed the experiments. ZG, JD and JJJ designed the methods. ZG, JD, JJJ, ZL, and XJH carried out all the statistical, computational analyses and drafted the manuscript. ZG, YH and SSL advised on all clinical aspects and interpreted the data. JD, JJJ, and XJH provided a critical review of the manuscript. All the authors have read the manuscript and approved the final version of the manuscript.

## Ethics Declarations

This study was approved by the Ethical Board of Tongji Medical College, Huazhong University of Science and Technology, China (Ethics code: UHCT-IEC-SOP-016-03-01).

## References

1. Joly, B. S., Coppo, P. & Veyradier, A. Thrombotic thrombocytopenic purpura. Blood 129, 2836–2846 (2017).

2. Mancini, I. et al. The HLA Variant rs6903608 Is Associated with Disease Onset and Relapse of Immune-Mediated Thrombotic Thrombocytopenic Purpura in Caucasians. J. Clin. Med. 9, (2020).

3. Sukumar, S. et al. Thrombotic Thrombocytopenic Purpura: Pathophysiology, Diagnosis, and Management. J. Clin. Med. 10, (2021).

4. Favaloro, E. J., Chapman, K., Mohammed, S., Vong, R. & Pasalic, L. Identification of ADAMTS13 Inhibitors in Acquired TTP. in Hemostasis and Thrombosis: Methods and Protocols (eds Favaloro, E. J. & Gosselin, R. C.) 505–521 (Springer US, New York, NY, 2023). doi:10.1007/978-1-0716-3175-1_33.

5. Lancellotti, S. et al. Immune and Hereditary Thrombotic Thrombocytopenic Purpura: Can ADAMTS13 Deficiency Alone Explain the Different Clinical Phenotypes? J. Clin. Med. 12, (2023).

6. Pishko, A. M., Li, A. & Cuker, A. Immune Thrombotic Thrombocytopenic Purpura: A Review. JAMA 334, 517–529 (2025).

7. Mancini, I. et al. Immunochip analysis identifies novel susceptibility loci in the human leukocyte antigen region for acquired thrombotic thrombocytopenic purpura. J. Thromb. Haemost. 14, 2356–2367 (2016).

8. Hrdinová, J. et al. Dissecting the pathophysiology of immune thrombotic thrombocytopenic purpura: interplay between genes and environmental triggers. Haematologica 103, 1099–1109 (2018).

9. Sakai, K. et al. HLA loci predisposing to immune TTP in Japanese: potential role of the shared ADAMTS13 peptide bound to different HLA-DR. Blood 135, 2413–2419 (2020).

10. Stubbs, M. J. et al. Identification of a novel genetic locus associated with immune-mediated thrombotic thrombocytopenic purpura. Haematologica 107, 574–582 (2022).

11. Thrombosis and Hemostasis Group, Chinese Society of Hematology, Chinese Medical Association. Chinese guideline on the diagnosis and management of thrombotic thrombocytopenic purpura. Chin. J. Hematol. 43, 7–12 (2022).

12. Joly, B. S., Coppo, P. & Veyradier, A. An update on pathogenesis and diagnosis of thrombotic thrombocytopenic purpura. Expert Rev. Hematol. 12, 383–395 (2019).

13. Chang, C. C. et al. Second-generation PLINK: rising to the challenge of larger and richer datasets. Gigascience 4, s13742–015-0047–8 (2015).

14. Turner, S. D. qqman: an R package for visualizing GWAS results using Q-Q and manhattan plots. J. Open Source Softw. 3, 731 (2018).

15. Willer, C. J., Li, Y. & Abecasis, G. R. METAL: fast and efficient meta-analysis of genomewide association scans. Bioinformatics 26, 2190–2191 (2010).

16. Zou, Y., Carbonetto, P., Wang, G. & Stephens, M. Fine-mapping from summary data with the “Sum of Single Effects” model. PLOS Genet. 18, e1010299 (2022).

17. Wang, G., Sarkar, A., Carbonetto, P. & Stephens, M. A Simple New Approach to Variable Selection in Regression, with Application to Genetic Fine Mapping. J. R. Stat. Soc. Ser. B Stat. Methodol. 82, 1273–1300 (2020).

18. McCreight, A. et al. SuSiE 2.0: improved methods and implementations for genetic fine-mapping and phenotype prediction. 2025.11.25.690514 Preprint at 10.1101/2025.11.25.690514 (2025).

19. Yuan, K. et al. Fine-mapping across diverse ancestries drives the discovery of putative causal variants underlying human complex traits and diseases. Nat. Genet. 56, 1841–1850 (2024).

20. Luo, Y. et al. A high-resolution HLA reference panel capturing global population diversity enables multi-ancestry fine-mapping in HIV host response. Nat. Genet. 53, 1504–1516 (2021).

21. Schaid, D. J., Rowland, C. M., Tines, D. E., Jacobson, R. M. & Poland, G. A. Score Tests for Association between Traits and Haplotypes when Linkage Phase Is Ambiguous. Am. J. Hum. Genet. 70, 425–434 (2002).

22. Joly, B. S. et al. HLA-DRB1*11 is a strong risk factor for acquired thrombotic thrombocytopenic purpura in children. Haematologica 105, e531–e534 (2020).

23. Coppo, P. et al. HLA-DRB1*11: a strong risk factor for acquired severe ADAMTS13 deficiency-related idiopathic thrombotic thrombocytopenic purpura in Caucasians. J. Thromb. Haemost. 8, 856–859 (2010).

24. Scully, M. et al. Human leukocyte antigen association in idiopathic thrombotic thrombocytopenic purpura: evidence for an immunogenetic link. J. Thromb. Haemost. 8, 257–262 (2010).

25. Hrdinová, J. et al. Mass spectrometry-assisted identification of ADAMTS13-derived peptides presented on HLA-DR and HLA-DQ. Haematologica 103, 1083– 1092 (2018).

26. Shiina, T., Hosomichi, K., Inoko, H. & Kulski, J. K. The HLA genomic loci map: expression, interaction, diversity and disease. J. Hum. Genet. 54, 15–39 (2009).

27. DeYoung, V., Singh, K. & Kretz, C. A. Mechanisms of ADAMTS13 regulation. J. Thromb. Haemost. 20, 2722–2732 (2022).

28. Hrdinová, J. et al. Dissecting the pathophysiology of immune thrombotic thrombocytopenic purpura: interplay between genes and environmental triggers. Haematologica 103, 1099–1109 (2018).

29. Brown, S. D. & Holt, R. A. Neoantigen characteristics in the context of the complete predicted MHC class I self-immunopeptidome. OncoImmunology 8, 1556080 (2019).

30. Laghmouchi, A., Graça, N. A. G. & Voorberg, J. Emerging Concepts in Immune Thrombotic Thrombocytopenic Purpura. Front. Immunol. 12, (2021).

31. Yang, L. et al. LINC00221 silencing prevents the progression of hepatocellular carcinoma through let-7a-5p-targeted inhibition of MMP11. Cancer Cell Int. 21, 202 (2021).

32. Huang, M. et al. LINC00221 suppresses the malignancy of children acute lymphoblastic leukemia. Biosci. Rep. 40, BSR20194070 (2020).

33. Feng, Y. et al. Clinical Value and Potential Mechanisms of LINC00221 in Hepatocellular Carcinoma Based on Integrated Analysis. Epigenomics 13, 299–317 (2021).

34. Chen, Z. et al. A construction and comprehensive analysis of the immune-related core ceRNA network and infiltrating immune cells in peripheral arterial occlusive disease. Front. Genet. 13, (2022).

35. Avnir, Y. et al. IGHV1-69 polymorphism modulates anti-influenza antibody repertoires, correlates with IGHV utilization shifts and varies by ethnicity. Sci. Rep. 6, 20842 (2016).

36. Olin, A. et al. Demographic and genetic factors shape the epitope specificity of the human antibody repertoire against viruses. 2023.11.07.23298153 Preprint at 10.1101/2023.11.07.23298153 (2024).

37. Lv, F. et al. Acute Hepatitis E Induced the First Episode of Immune-Mediated Thrombotic Thrombocytopenic Purpura: The First Case Report. Infect. Drug Resist. 16, 5149–5154 (2023).

38. Amitai, A. et al. Defining and Manipulating B Cell Immunodominance Hierarchies to Elicit Broadly Neutralizing Antibody Responses against Influenza Virus. Cell Syst. 11, 573–588.e9 (2020).

39. Ataca, S. et al. Modulating the immunodominance hierarchy of immunoglobulin germline-encoded structural motifs targeting the influenza hemagglutinin stem. Cell Rep. 43, 114990 (2024).

40. Hwang, K.-K. et al. IGHV1-69 B Cell Chronic Lymphocytic Leukemia Antibodies Cross-React with HIV-1 and Hepatitis C Virus Antigens as Well as Intestinal Commensal Bacteria. PLOS ONE 9, e90725 (2014).

41. Teo, Q. W. et al. Stringent and complex sequence constraints of an IGHV1-69 broadly neutralizing antibody to influenza HA stem. Cell Rep. 42, 113410 (2023).

42. Hao, H. et al. FUT10 and FUT11 are protein O-fucosyltransferases that modify protein EMI domains. Nat. Chem. Biol. 21, 598–610 (2025).

43. Žárský, V. & Doležal, P. Evolution of the Tim17 protein family. Biol. Direct 11, 54 (2016).

44. Guy, A. et al. Vascular endothelial cell expression of JAK2V617F is sufficient to promote a pro-thrombotic state due to increased P-selectin expression. Haematologica 104, 70–81 (2019).

45. Prajapat, S. K., Maharana, K. Ch. & Singh, S. Mitochondrial dysfunction in the pathogenesis of endothelial dysfunction. Mol. Cell. Biochem. 479, 1999–2016 (2024).

46. Johnson, C. F. et al. RIPK3 Protects Against Endothelial Activation and Vascular Permeability in a Mouse Model of Ischemia-Reperfusion Injury. Arterioscler. Thromb. Vasc. Biol. 46, 148–164 (2026).

47. Kumar, R. et al. Oligonucleotide correction of an intronic TIMMDC1 variant in cells of patients with severe neurodegenerative disorder. Npj Genomic Med. 7, 9 (2022).

48. Bonnez, Q., Sakai, K. & Vanhoorelbeke, K. ADAMTS13 and Non-ADAMTS13 Biomarkers in Immune-Mediated Thrombotic Thrombocytopenic Purpura. J. Clin. Med. 12, (2023).

49. Incalza, M. A. et al. Oxidative stress and reactive oxygen species in endothelial dysfunction associated with cardiovascular and metabolic diseases. Vascul. Pharmacol. 100, 1–19 (2018).

50. Higashi, Y. Roles of Oxidative Stress and Inflammation in Vascular Endothelial Dysfunction-Related Disease. Antioxidants 11, 1958 (2022).

