## Supplementary files for "Genetic architecture of immune-mediated thrombotic thrombocytopenic purpura in Han Chinese individuals"

**Genome-wide association study provides novel functional insights into thrombotic thrombocytopenic purpura in Han Chinese population**

**Supplementary Tables**

Table S1. Characteristics of the discovery and validation cohorts.

Table S2. Suggestive association signals for thrombotic thrombocytopenic purpura.

Table S3. Fine-mapping credible-set variants at the 6p21 and 14q32 loci.

Table S4. Association signals before and after conditioning on HLA-DRB1 amino-acid position 74-LRx.

Table S5. Interaction analysis between AA-DRB1-74-LRx and HLA-DRB1*08:03.

**Supplementary Figures**

Figure S1. Quality-control diagnostics for the discovery cohort.

Figure S2. Quality-control diagnostics for the validation cohort.

Figure S3. Genome-wide association results in the discovery cohort.

Figure S4. Genome-wide association results in the validation cohort.

Figure S5. Conditional association analysis of the MHC region.

Figure S6. Consistency of LD correlation between HLA-DRB1 AA74 and classical HLA alleles.

Figure S7. Regional association plots for lead GWAS loci.

Figure S8. SuSiE-RSS fine-mapping posterior inclusion probabilities at the 6p21 and 14q32 loci.

**Supplementary Tables**

**Table S1. Characteristics of the discovery and validation cohorts.**

| Cohort | Total | Cases | Controls | Age | Male | Female | Variants after QC |
| --- | --- | --- | --- | --- | --- | --- | --- |
| Discovery | 913 | 174 | 743 | NA | 471 (51.4%) | 446 (48.6%) | 524,866 |
| Validation | 431 | 129 | 302 | 46.0 [35.0, 57.0] | 189 (43.9%) | 242 (56.1%) | 510,182 |

Age is shown as median [interquartile range]. Age information was available for the validation cohort only. Sex is shown as number and percentage. SNPs after QC indicate the number of autosomal variants retained after genotype quality control. Abbreviations: IQR, interquartile range; NA, not available; QC, quality control; SNP, single nucleotide polymorphism; TTP, thrombotic thrombocytopenic purpura.

**Table S2. Suggestive significant variant associated with thrombotic thrombocytopenic purpura**

| rsID | Chr | Position (Hg19) | Risk Allele | Non-risk Allele | Stage | Risk allele frequency  TTP Control | | OR (95% CI) | *P*-value |
| --- | --- | --- | --- | --- | --- | --- | --- | --- | --- |
| rs114311228 | 4 | 188946168 | A | G | Discovery | 0.94 | 0.989 | 0.18 (0.09-0.35) | 9.26e-07 |
|  |  |  |  |  | Validation | 0.965 | 0.987 | 0.41 (0.14-1.24) | 1.14e-01 |
|  |  |  |  |  | Meta-Analysis | 0.951 | 0.988 | 0.22 (0.13-0.40) | 6.01e-07 |
| rs6930615 | 6 | 32392205 | A | G | Discovery | 0.15 | 0.054 | 3.12 (2.00-4.88) | 5.48e-07 |
|  |  |  |  |  | Validation | 0.106 | 0.064 | 1.55 (0.85-2.82) | 1.55e-01 |
|  |  |  |  |  | Meta-Analysis | 0.131 | 0.057 | 2.43 (1.70-3.48) | 1.12e-06 |
| rs9843355 | 3 | 119228508 | A | G | Discovery | 0.224 | 0.356 | 0.53 (0.40-0.69) | 4.78e-06 |
|  |  |  |  |  | Validation | 0.264 | 0.339 | 0.67 (0.47-0.95) | 2.30e-02 |
|  |  |  |  |  | Meta-Analysis | 0.241 | 0.351 | 0.58 (0.47-0.72) | 5.93e-07 |

Positions are based on GRCh37/hg19. The effect allele is the allele tested in the association model, and effect-allele frequencies are shown separately for TTP cases and controls. Odds ratios and 95% confidence intervals are reported for the effect allele. The suggestive significance threshold was calculated after LD pruning, which identified 232,189 independent SNPs; therefore, the suggestive threshold was set at *P* = 4.31 × 10^−6^. Abbreviations: Chr, chromosome; CI, confidence interval; LD, linkage disequilibrium; OR, odds ratio; TTP, thrombotic thrombocytopenic purpura.

**Table S3. Fine-mapping credible-set variants at the 6p21 and 14q32 loci.**

| rsID | Chr | Position (Hg19) | Ref | Alt | Beta | SE | *P*-value | | PIP | r^2^ to lead |
| --- | --- | --- | --- | --- | --- | --- | --- | --- | --- | --- |
| rs2187668 | 6 | 32605884 | G | A | 1.3722 | 0.221 | | 5.348e-10 | 0.978187837 | 1 |
| rs1024350 | 14 | 107141122 | G | A | 0.64 | 0.099 | | 1.006e-10 | 0.804648434 | 1 |
| GSA-rs7157975 | 14 | 107212287 | G | A | 0.6005 | 0.0992 | | 1.423e-09 | 0.155331793 | 0.912609442 |

Positions are based on GRCh37/hg19. Beta and SE are from the association summary statistics used for SuSiE-RSS fine-mapping. PIP indicates posterior inclusion probability. r² to lead indicates linkage disequilibrium with the lead variant within each locus. Abbreviations: Chr, chromosome; PIP, posterior inclusion probability; SE, standard error; SuSiE-RSS, Sum of Single Effects regression using summary statistics.

**Table S4. Association signals before and after conditioning on HLA-DRB1 amino-acid position 74-LRx.**

| Variants | *P*-value (original) | Direction  (original) | *P*-value (conditional) | Direction (conditional) |
| --- | --- | --- | --- | --- |
| rs6906021 | 1.974e-06 | -- | 0.00142 | -- |
| rs2187668 | 1.833e-05 | ++ | 0.1001 | ++ |
| rs6929590 | 3.26e-05 | -- | 0.5739 | -+ |
| rs7753500 | 3.26e-05 | -- | 0.5738 | -+ |
| rs6457580 | 3.262e-05 | ++ | 0.5741 | -+ |
| rs114656303 | 3.263e-05 | -- | 0.5742 | -+ |
| rs16822586 | 3.263e-05 | ++ | 0.5742 | -+ |
| rs6911791 | 3.263e-05 | ++ | 0.5742 | -+ |

Conditional association analysis was performed by adjusting for the dosage of AA-DRB1-74-LRx. The original P value refers to the association before conditioning, and the conditional P value refers to the association after adjustment for AA-DRB1-74-LRx. Direction indicates the effect direction across cohorts, with signs shown in cohort order.

**Table S5. Interaction analysis between AA-DRB1-74-LRx and HLA-DRB1*08:03.**

| Interaction Variable | Estimate | Std. Error | z value | *P*-value |
| --- | --- | --- | --- | --- |
| AA-DRB1-74-LRx + HLA-DRB1*08:03 | 0.289 | 0.42 | 0.68 | 0.50 |

The estimate represents the interaction term between AA-DRB1-74-LRx and HLA-DRB108:03 in the logistic regression model. The non-significant interaction P value indicates no statistical evidence that the effect of AA-DRB1-74-LRx differs according to HLA-DRB108:03 carrier status. Abbreviation: SE, standard error.

**Supplementary Figures**

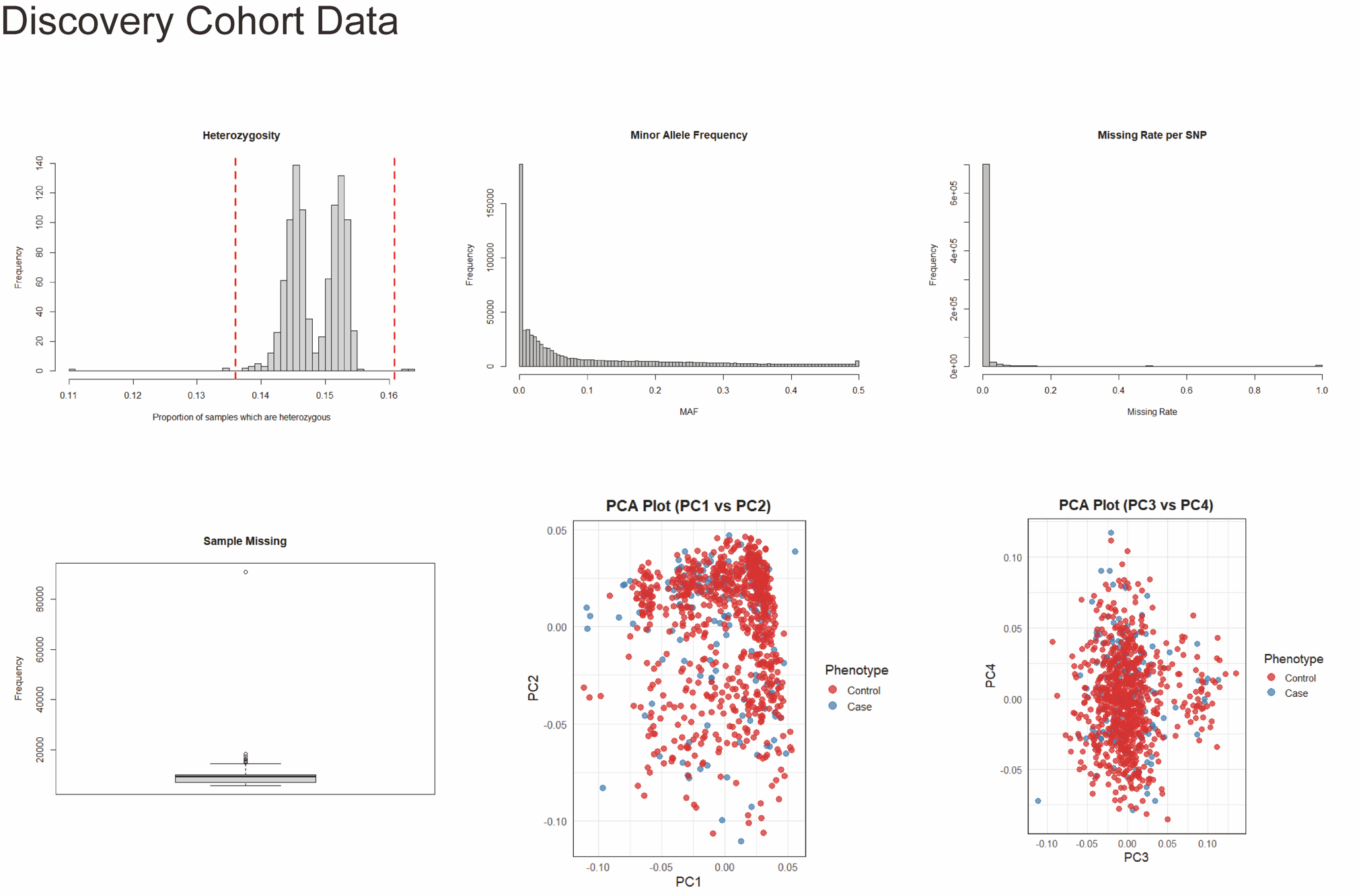

**Figure S1. Quality-control diagnostics for the discovery cohort.**

Distributions of heterozygosity, minor allele frequency, SNP missingness, and sample missingness are shown for the discovery cohort, together with principal component analysis plots for PC1 versus PC2 and PC3 versus PC4. Cases and controls are shown separately to evaluate genotype quality and population structure before downstream association analysis.

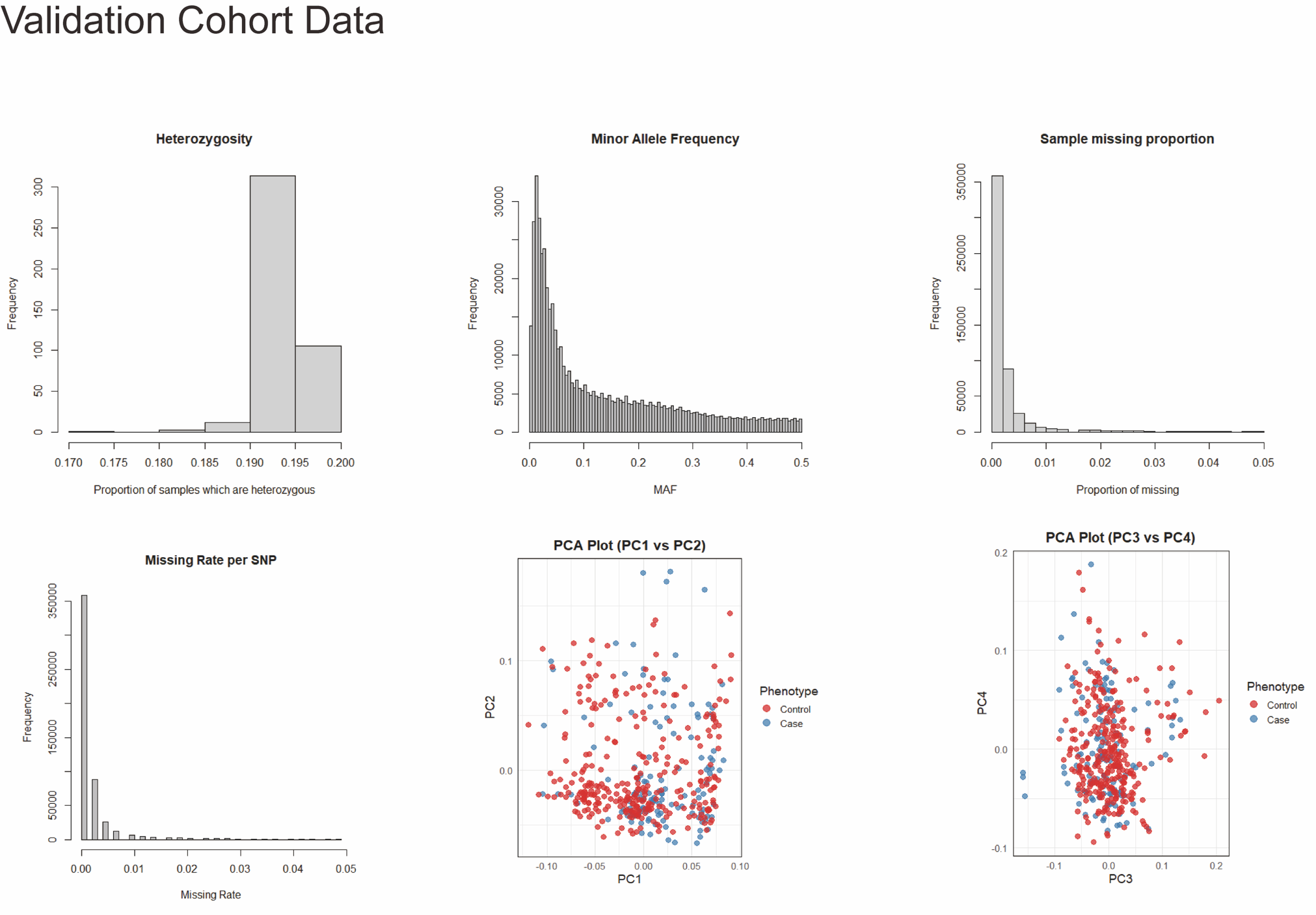

**Figure S2. Quality-control diagnostics for the validation cohort.**

Distributions of heterozygosity, minor allele frequency, sample missing proportion, and SNP missingness are shown for the validation cohort, together with PCA plots for PC1 versus PC2 and PC3 versus PC4. These diagnostic plots were used to confirm sample quality and ancestry clustering before association testing.

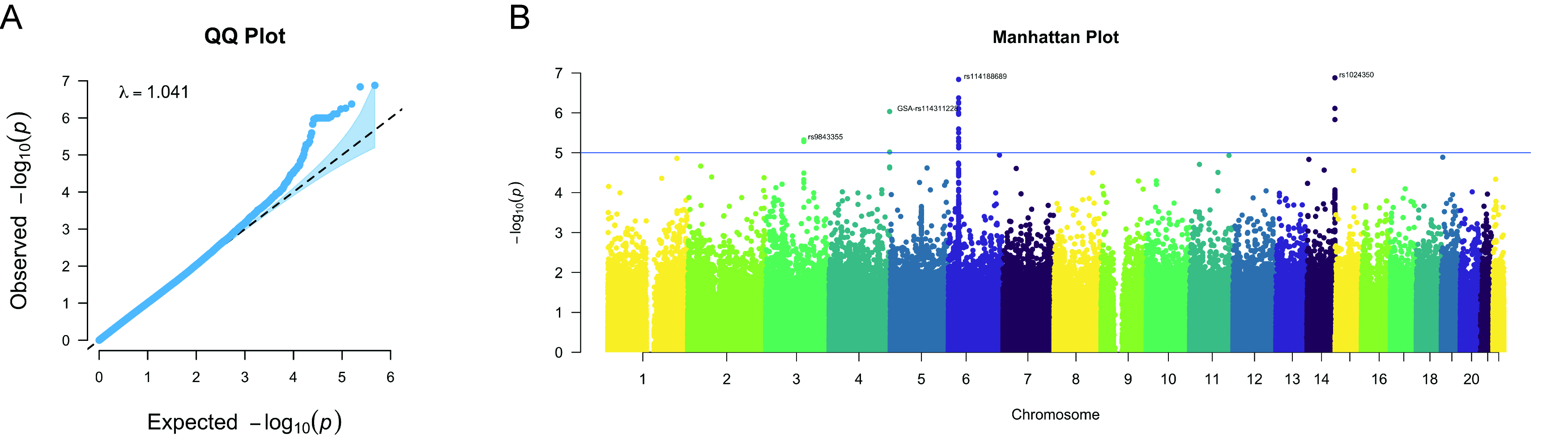

**Figure S3. Genome-wide association results in the discovery cohort.**

(A) Quantile-quantile plot of association P values in the discovery cohort, showing limited genomic inflation. (B) Manhattan plot of genome-wide association results across autosomes. The horizontal reference line indicates the suggestive significance threshold.

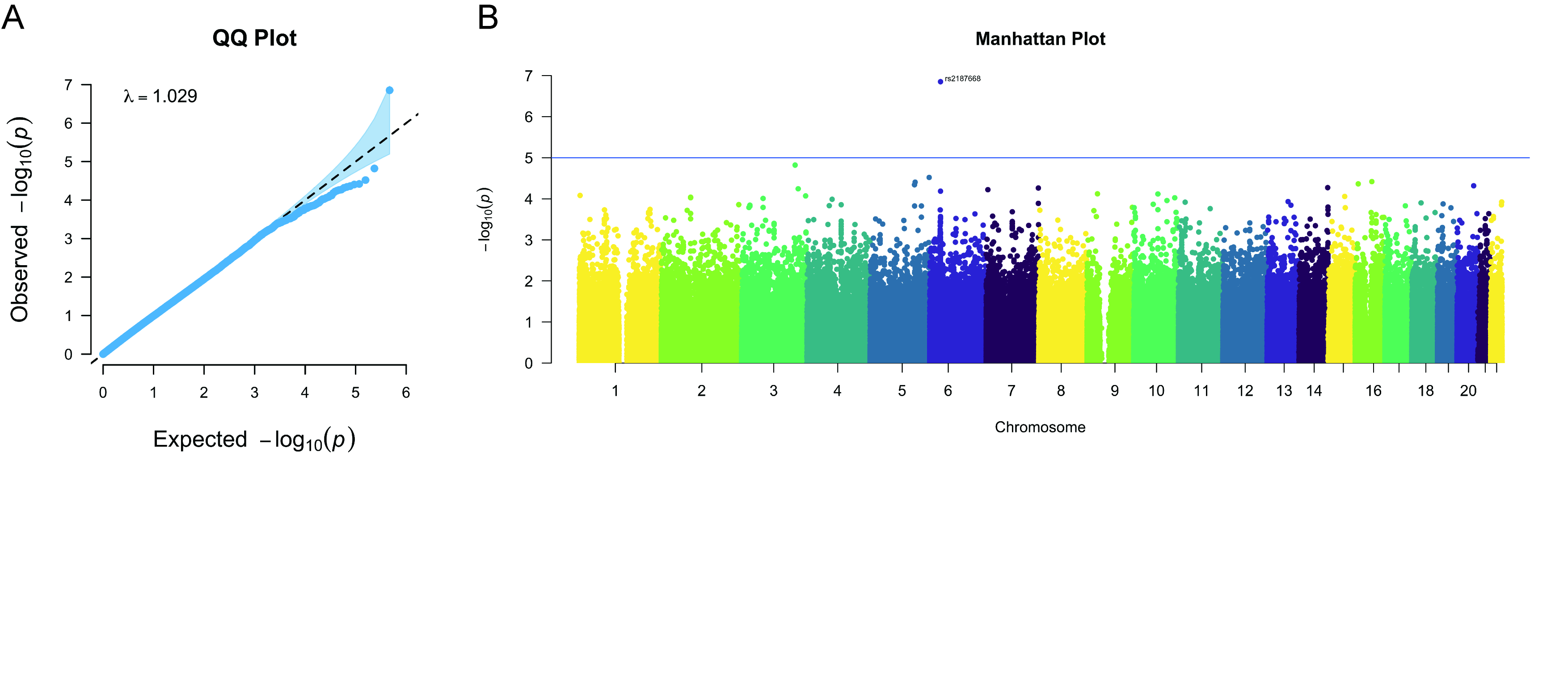

**Figure S4. Genome-wide association results in the validation cohort.**

(A) Quantile-quantile plot of association P values in the validation cohort. (B) Manhattan plot of association signals across autosomes in the validation cohort. The genomic inflation factor indicates that population stratification was adequately controlled.

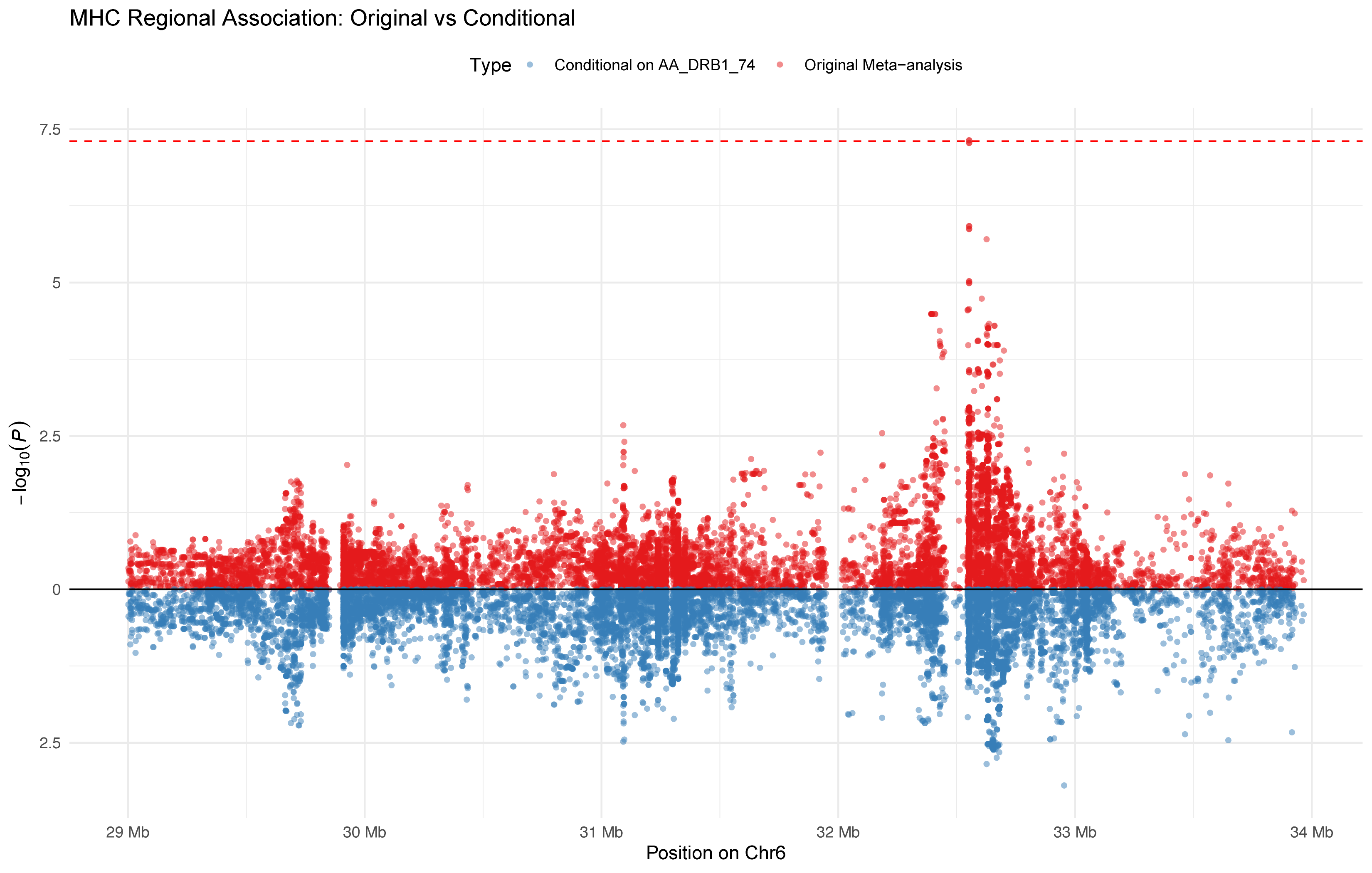

**Figure S5. Conditional association analysis of the MHC region.**

Regional association signals across the MHC region before and after conditioning on HLA-DRB1 amino-acid position 74 are shown. The marked attenuation of association signals after conditioning supports AA-DRB1-74 as the principal driver of the MHC association with TTP susceptibility.

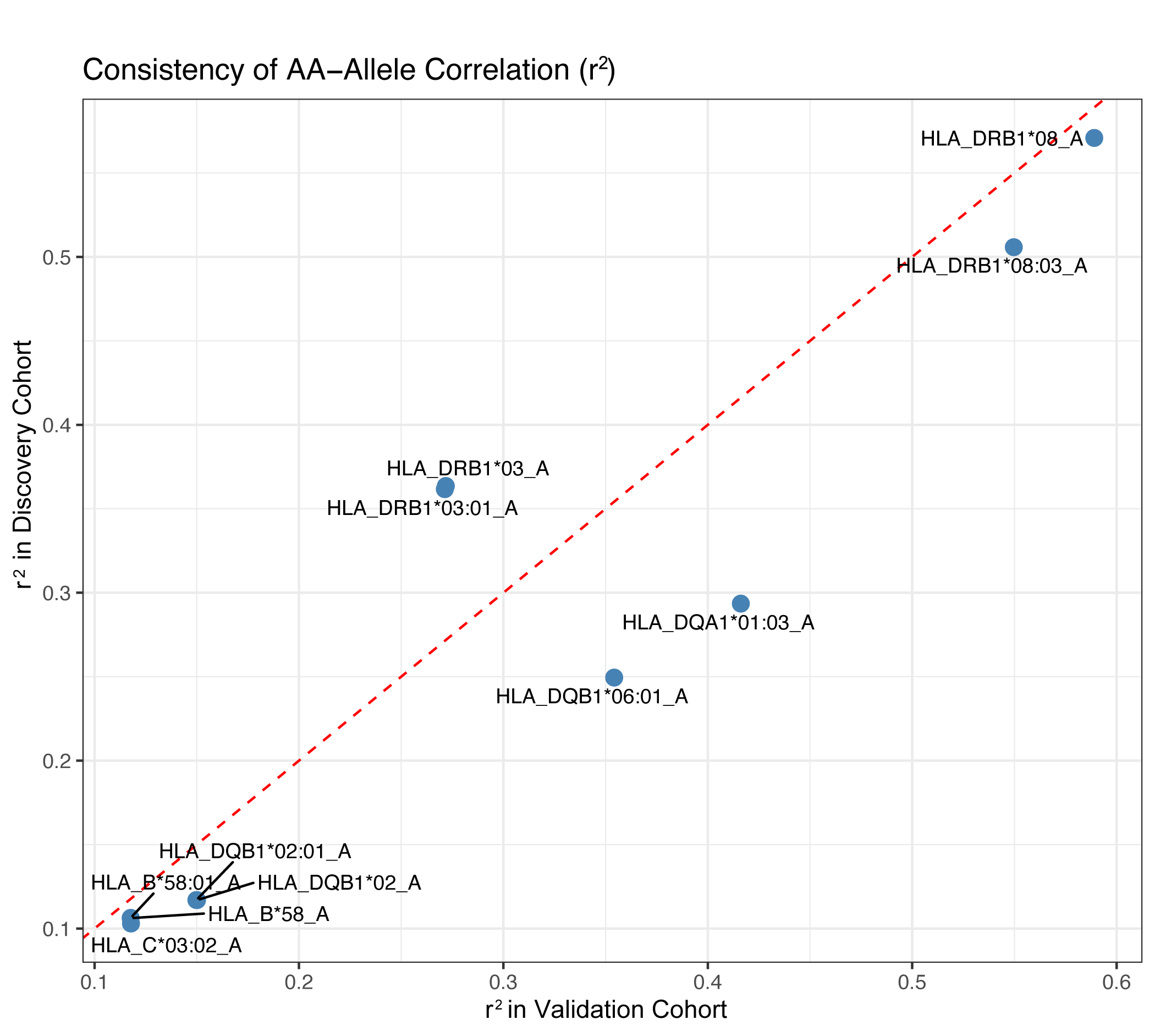

**Figure S6. Consistency of LD correlation between HLA-DRB1 AA74 and classical HLA alleles.**

The scatter plot compares linkage disequilibrium correlation values between HLA-DRB1 AA74 and classical HLA alleles in the discovery and validation cohorts. The diagonal reference line indicates concordance between cohorts, supporting consistent AA-allele correlation patterns.

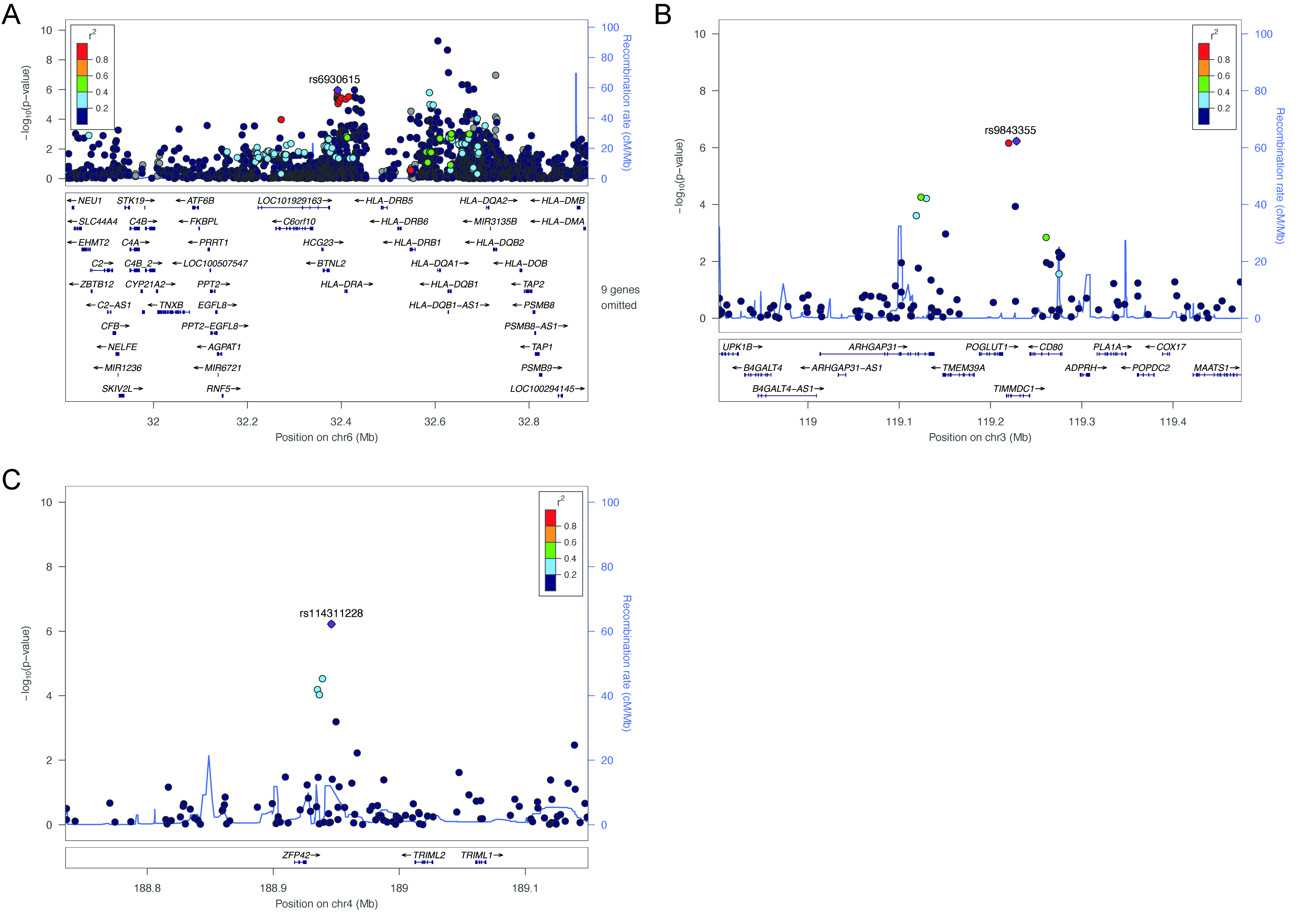

**Figure S7. Regional association plots for lead GWAS loci.**

LocusZoom plots are shown for the major association regions identified in the GWAS meta-analysis, including the MHC locus on chromosome 6, the chromosome 3 signal, and the chromosome 14 locus. Points are colored according to linkage disequilibrium with the lead variant, and recombination rates are shown in blue.

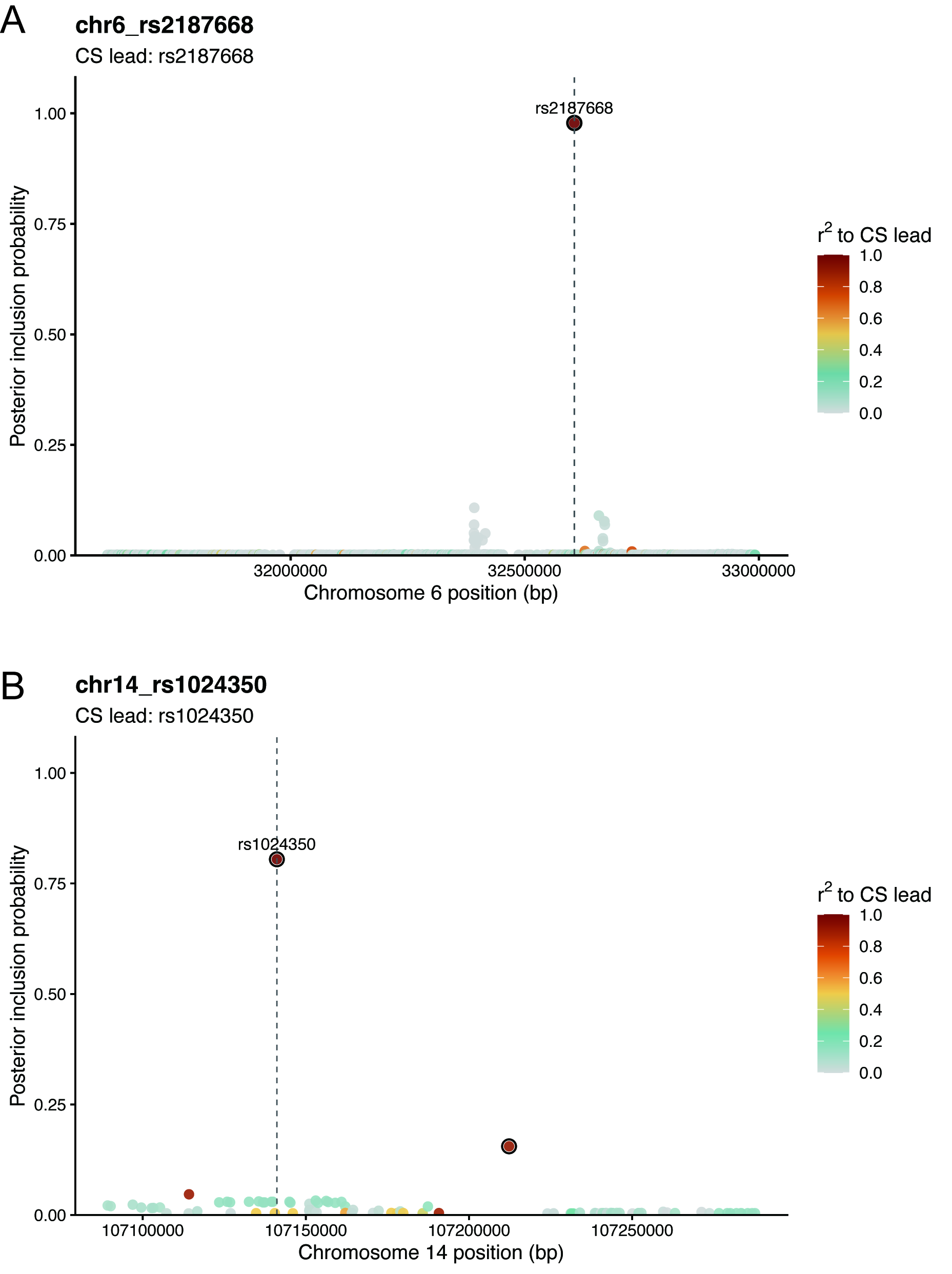

**Figure S8. SuSiE-RSS fine-mapping posterior inclusion probabilities at the 6p21 and 14q32 loci.**

Posterior inclusion probabilities are plotted by genomic position. Points are colored according to linkage disequilibrium (r2) with the credible-set lead variant, and dashed vertical lines indicate the lead variants. Fine-mapping prioritized rs2187668 at 6p21 and rs1024350 at 14q32.
